# Reducing Stigma Among Providers Caring for Pregnant Patients with Substance Use Disorders: A Systematic Review of Interventions

**DOI:** 10.1101/2025.06.17.25329533

**Authors:** Karli S Swenson, Sydney Comstock, Sarah Briley

## Abstract

**Background:** The prevalence of substance use disorders (SUDs) among pregnant individuals has risen alongside the opioid epidemic, contributing to increased maternal morbidity and mortality. Many pregnant individuals with SUDs experience significant stigma and bias from the healthcare system, which discourages them from seeking necessary care. This stigma, often exacerbated by fears of child protective services involvement, can prevent patients from engaging in treatment, thereby impacting maternal and fetal health. Reducing stigma among healthcare providers and nurses is critical to improving care and outcomes for this population. This systematic review aims to identify and compile primary literature on interventions designed to decrease stigma in providers and nurses caring for pregnant individuals with SUDs.

**Methods:** We conducted a systematic search of Google Scholar, Web of Science, and Embase using comprehensive terms related to substance use, healthcare providers, stigma, and pregnancy. Only primary research articles were included, with exclusions for review papers, meta-analyses, and commentaries, as well as studies focused on unrelated topics (e.g., prescriptions, schizophrenia, psychosis). Using Covidence software, we screened 2,308 articles, with 558 duplicates removed automatically. Double-blind title and abstract screening resulted in the exclusion of 1,330 articles, leaving 420 for full-text review. After applying inclusion criteria, 19 studies were included in the final analysis.

**Results:** The 19 included studies represented a wide range of educational interventions designed to reduce provider stigma toward pregnant individuals with SUDs. Interventions included online learning modules, professional development workshops, clinical immersion experiences, and one arts-based program. While training formats and evaluation tools varied widely, most studies reported improvements in provider knowledge, confidence, and attitudes. However, fewer demonstrated sustained behavior change, and only a minority used validated instruments or long-term follow-up.

**Conclusions:** Stigma reduction interventions for providers caring for pregnant people with SUDs are becoming more common, particularly in response to rising perinatal substance use rates. Despite the effectiveness of many approaches, intervention and evaluation strategies remain non-standardized. Expanding access to training, especially in high-burden and under-resourced settings, and developing validated, scalable, and emotionally engaging education models will be critical to improving perinatal care quality and equity.

## INTRODUCTION

Substance use disorders (SUDs) among pregnant individuals have become an escalating public health concern, paralleling the rise of the opioid epidemic in the United States^1^. The rates of substance use among pregnant individuals have significantly increased, leading to profound consequences on maternal and fetal health.^1^ In Colorado, accidental overdose has emerged as the second leading cause of death among pregnant and postpartum individuals, underscoring the critical need for effective interventions and support within this population.^2^ Beyond mortality, substance use contributes to severe maternal morbidity, especially when compounded by co-occurring mental health conditions, socioeconomic stressors, and systemic racial biases that hinder access to comprehensive care.^3^

Many pregnant individuals with SUDs face substantial barriers to accessing healthcare, exacerbated by pervasive stigma and fear of repercussions, including potential involvement of child protective services (CPS).^4,5^ This stigma is often rooted in societal biases that characterize substance use in pregnancy as a moral failing rather than a medical condition, leading to severe judgment and alienation.^4^ Healthcare providers, including those on labor and delivery, postpartum, nursery, and neonatal intensive care unit (NICU) teams, may consciously or unconsciously express these biases, which can manifest as differential treatment, judgmental comments, or less compassionate care. These attitudes often leave patients feeling degraded, unworthy of care, or afraid to disclose substance use, ultimately restricting their access to vital medical support.^4^

The fear of negative treatment or punitive measures deters many from disclosing their substance use to their obstetrician, midwife, nurse, or other healthcare providers, limiting access to necessary treatment and support during a vulnerable period.^6^ Compounded by racial and socioeconomic biases that already affect the quality of care for many marginalized individuals, the stigma surrounding SUDs in pregnancy isolates patients from health resources and heightens their risks of adverse health outcomes.^7^ Addressing these deeply embedded biases is critical to fostering a healthcare environment where all pregnant individuals feel safe seeking treatment for substance use, therefore receiving necessary care.

An essential, shared goal between patients and healthcare providers should be to foster a supportive environment where pregnant individuals with SUDs feel encouraged to seek and maintain treatment. Key objectives in achieving this include promoting access to and understanding of medication for opioid use disorder (MOUD) or other pharmacologic treatments, facilitating honest communication with healthcare providers, and supporting engagement in critical healthcare appointments, whether obstetric, postpartum, pediatric, or mental health services. Pregnant patients must experience positive, non-judgmental, and transparent care interactions, underscoring the need to reduce stigma and bias among healthcare providers.

This systematic review aims to examine the current evidence on how to decrease healthcare provider stigma and bias toward pregnant individuals with SUDs and to explore strategies that can enhance ethical and effective care in obstetric settings.

## METHODS

This systematic review aimed to compile primary literature examining methods to decrease stigma among healthcare providers caring for pregnant individuals with SUDs. We included only primary research articles and excluded any reviews, meta-analyses, or commentaries to maintain a focus on original research findings.

### Search Strategy

We conducted a comprehensive literature search across three scientific databases: Google Scholar, Web of Science, and Embase. The search strategy was designed to capture studies examining healthcare provider stigma related to substance use during pregnancy. Search terms included keywords and variations relevant to four primary domains: substance use (e.g., “substance*,” “substance use disorder,” “drug abuse,” “alcohol dependence,” “opioid,” “addict*”), healthcare providers (e.g., “provider*,” “clinician*,” “nurs*,” “therapist*,” “lactation consultant”), stigma (e.g., “stigma*,” “attitude*,” “bias*,” “discriminat*”), and pregnancy (e.g., “pregnan*,” “fetus,” “gestation*”). Boolean operators were used to combine terms and refine the search. To maintain focus on the population of interest and the healthcare context, we applied exclusion criteria to omit studies primarily focused on unrelated conditions or populations, including “prescription,” “schizophrenia,” “antiretroviral,” “psychosis,” “obesity,” “epilepsy,” “tuberculosis,” and “hepatitis.”

### Screening and Selection

Using Covidence software, which is designed for comprehensive literature reviews, we imported and screened 2,308 manuscripts retrieved from the database search. Covidence detected and auto-removed 558 duplicate records, leaving 1,750 unique studies. Two independent reviewers conducted a double-blind screening of titles and abstracts to assess relevance to the study aim, with 1,330 studies deemed irrelevant at this stage. Conflicts between two reviewers were resolved by the lead reviewer. Full-text reviews were conducted on the remaining 420 studies, of which 401 were excluded based on predetermined inclusion criteria related to the study population, outcomes, and intervention focus. Ultimately, 19 studies met all inclusion criteria and were included in the final analysis.

### Data extraction

Data extracted from the manuscripts included title, year of publication, location, lead author contact information, methods, audience for the training intervention, substance focus for the intervention, outcomes from the intervention, and the method of measuring the outcomes.

## RESULTS

Of the 19 included studies, 16 were conducted in the United States, two in Australia and one in the United Kingdom, and were published between 2007 and 2024. 11 studies focused on substance use in broad terms, whereas four focused specifically on alcohol use and four on opioid use. The most common methodologies were professional education webinars (12 studies) that ranged from 20 minutes to 4 hours, followed by online resource modules (3 studies) and clinical immersion programs (3 studies), followed by one study that did a feasibility analysis for an arts-based stigma intervention program.

### Professional education webinars

A substantial subset of the included studies (n=12) focused on professional development training, typically delivered through structured workshops, in-person or hybrid learning sessions, or formal continuing education curricula. These interventions shared common goals: to increase provider knowledge, shift attitudes, and improve confidence and communication skills when caring for pregnant individuals who use substances.

#### Intervention Similarities and Differences

Many of these interventions incorporated pre-post survey designs alongside didactic content, case studies, or patient narratives to encourage reflection and build empathy. For example, Zoorob^8^ (2014, n=420) and Wright^9^ (2022, n=42) provided targeted content on fetal alcohol spectrum disorders or opioid use disorder, respectively, using brief, evidence-based training sessions focused on improving confidence and clinical identification skills. Seybold^10^ (2014, n=70) and Howard^11^ (2017, n=29) emphasized interdisciplinary, competency-based workshops, incorporating trauma-informed care principles, child welfare context, and recovery-oriented communication. A few studies also embedded interactive, interprofessional components. West^12^ (2022, n=104) used a hybrid training model combining online modules with a one-day in-person event focused on cross-agency communication and collaboration for families affected by substance use. Hooks^13^ (2019, n=48) uniquely integrated peer discussion boards and semi-structured interviews into their educational strategy, helping final-year midwifery students reflect on stigma and bridge the “theory-practice divide.”

Roberts^14^ (2024, n=592) and Ford^15^ (2021, n=1,496) offered large-scale quality improvement training programs aiming to reduce providers overreporting to child welfare and reframe attitudes about mothers with opioid use disorder. These trainings were typically embedded into statewide or institutional quality initiatives, underscoring the potential for wide-scale implementation. Meanwhile, Merritt^16^ (2022, n=89) and Wingo^17^ (2023, n=42) used community-informed training approaches - Wingo in particular partnered with individuals with lived experience to co-develop the intervention. Finally, Taniguchi^18^ (2023, n=13) and Thomas^19^ (2024, n=128) offered focused sessions for perinatal nurses, aiming to improve knowledge and reduce bias using structured content on neonatal opioid withdrawal, care planning, and the ethics of substance use-related reporting.

#### Evaluation Approaches

Most of the studies relied on pre- and post-test surveys measuring changes in knowledge, attitudes, confidence, and/or intended practices. Some, like Merritt^16^ and Roberts^14^, used repeated measures designs, adding a follow-up survey (e.g., at 1 month or 6 months) to evaluate sustained change. Instruments varied widely, from established tools like the Attitudes of Healthcare Providers Survey (AHPS) and the Medical Condition Regard Scale, to custom or adapted Likert-scale questionnaires. Several studies also included open-ended responses or qualitative components (e.g., Hooks^13^, Wingo^17^, Seybold^10^) to better understand provider perceptions and reactions to the training content.

#### Effectiveness of Interventions

Across these studies, professional development training was generally effective, particularly in enhancing knowledge, confidence, and empathy toward patients. For example: Zoorob^8^, Wright^9^, and Taniguchi^18^ showed statistically significant increases in provider knowledge or confidence following brief trainings. Roberts^14^ and Ford^8^ demonstrated measurable reductions in punitive attitudes toward child welfare reporting and increased compassion toward mothers of infants with neonatal abstinence syndrome (NAS). Wingo^17^ found improved comfort with reproductive counseling and reduced bias toward parenting intentions of substance-using individuals. Hooks^13^ and Seybold^10^ highlighted the importance of empathy building and reflective practice, noting qualitative shifts in how participants conceptualized addiction and care. West^12^ and Howard^11^ emphasized collaborative communication strategies, with participants reporting enhanced interprofessional coordination. While sustainability of changes varied across studies, several (e.g., Merritt^16^, Roberts^14^) found that some effects waned over time, reinforcing the need for ongoing support, institutional culture change, and access to community-based resources to maintain gains.

### Online resource modules

Three studies - Payne 2011^20^ (n=1,001 health professionals), Payne 2011^21^ (n=82 pediatricians), and Kennedy-Hendricks 2022^22^ (n=1,842 healthcare providers) - implemented educational interventions that relied on online or distributed materials to improve healthcare provider knowledge, attitudes, and practices related to substance use in pregnancy. Despite differences in scope, location, and measurement, all three studies shared the common goal of using accessible, scalable materials to reduce stigma and increase provider competence.

The two Payne 2011 studies^20,21^, both conducted in Australia by the same investigators, involved the distribution of educational resources about fetal alcohol spectrum disorder (FASD) to health professionals (general and pediatric) and measured changes approximately six months after the materials were sent. The larger study included a wide range of professionals: nurses, general practitioners, allied health professionals, and obstetricians, while the second focused specifically on pediatricians. In both cases, participants received paper-based or web-based resources developed based on earlier surveys. These interventions were passive in delivery but had clear content tailored to clinical practice. In contrast, the Kennedy-Hendricks^22^ 2022 study, conducted in the United States, utilized randomized visual campaigns and narrative vignettes within a web-based format to examine the impact of different message framings and messengers on stigma toward individuals with opioid use disorder (OUD). This study stands out for its controlled design and message testing, incorporating both “Words Matter” (language-focused) and “Medication Treatment Works” (treatment-focused) campaigns, with narratives presented by simulated patients, clinicians, or administrators.

The evaluation approaches also varied notably. Both Payne^13,21^ studies used prevalence rate ratios and confidence intervals to compare survey results before and after the intervention, focusing on changes in knowledge, attitudes, and clinical practice (e.g., understanding features of FAS, willingness to advise alcohol abstinence, or frequency of screening). Kennedy-Hendricks^22^, by contrast, employed a randomized clinical trial design and analyzed changes in stigma using Likert-scale items and feeling thermometer scores for warmth (general feelings towards the person with SUD), social distance (willingness to engage in close social relationships with individuals with SUD), and blame (attribute personal responsibility to individuals with SUD). This methodological rigor allowed for more nuanced detection of causal effects from specific messaging strategies. All three studies demonstrated partial or full effectiveness. In the general Payne 2011 study^13^, 48.5% of respondents reported that the materials influenced or changed their clinical practice, and knowledge about FASD increased modestly.

Pediatricians in the second Payne 2011 paper^21^ showed increased endorsement of complete alcohol abstinence during pregnancy but no change in the proportion who routinely asked about alcohol use. This highlights a disconnect between knowledge/attitude shifts and behavioral practice change. Kennedy-Hendricks^22^ 2022 demonstrated strong, statistically significant reductions in stigma toward individuals with OUD, particularly when messages were paired with a patient narrative. Participants were less likely to prefer social distance and rated individuals with OUD with significantly higher warmth scores, illustrating the value of personalized, patient-centered storytelling in reducing stigma.

### Clinical Immersion Programs

Three studies, Meng^23^ (2007, n=117), Ramirez-Cacho^24^ (2007, n=104), and Albright^25^ (2012, n=96), used clinical immersion programs as interventions to reduce stigma and improve attitudes among medical students toward pregnant individuals with SUDs. All three studies integrated exposure to specialized prenatal care settings during obstetrics-gynecology rotations, offering medical students real-world opportunities to engage with patients experiencing substance use in pregnancy. While each study utilized a similar foundation, participation in a half-day prenatal clinic specifically designed for women with SUDs, the design diverged slightly. Albright^25^ built on the approach used by Ramirez-Cacho^24^ by not only including the prenatal clinic experience but also adding a randomized component where some students attended a rehabilitation residence for pregnant individuals. This second site deepened students’ understanding by exposing them to patients’ lived experiences with recovery, including barriers to care and efforts to set therapeutic goals. Meng^23^, in contrast, had a simpler design, where students were assigned to attend the clinic or not, without the additional exposure to a residential setting.

All three studies used pre- and mid-rotation or post-rotation Likert-scale surveys to assess changes in student attitudes and comfort levels. The evaluations in Meng^23^ and Ramirez-Cacho^24^ were similar in that they both used a 5-point scale assessing comfort with discussing substance use and attitudes toward patients. Albright^25^, while also using a Likert-style instrument, divided the survey into two clear domains: six items addressing comfort and six addressing attitudes, allowing for more nuanced analysis of change across different constructs. In terms of effectiveness, all three studies found positive outcomes. Meng^23^ reported increased comfort in inquiring about alcohol use and decreased beliefs associating addiction with weak will. Ramirez-Cacho^24^ found that students who attended the clinic reported being more nonjudgmental and comfortable discussing substance use, whereas the control group exhibited a decline in comfort and awareness over time. Albright^25^ showed that both the clinic and the residence experiences contributed to greater comfort discussing substance use and an increased understanding of the challenges faced by patients, with enhanced outcomes for those who attended both.

### Arts-based intervention

Shuman^26^ (2024, n=11) piloted a novel arts-based intervention called *ArtSpective™* designed to reduce stigma among healthcare trainees toward pregnant individuals with SUD. This proof-of-concept, mixed-methods quasi-experimental study was conducted with undergraduate and graduate nursing students who had prior experience in maternal-infant care. The intervention utilized immersive artistic media to engage participants emotionally and cognitively, aiming to foster empathy and shift stigmatizing attitudes. Participants completed pre- and post-intervention surveys, including an adapted version of the Attitudes About Drug Use in Pregnancy Scale, along with qualitative feedback collected through a focus group. The quantitative findings showed a statistically significant reduction in stigma (p = 0.003; Cohen’s d = 0.633), suggesting a moderate effect size. Qualitatively, participants expressed high satisfaction with the intervention and emphasized its potential scalability and relevance.

## DISCUSSION

This review identified 19 studies evaluating interventions to reduce stigma and bias among healthcare providers caring for pregnant individuals with SUD. The number of publications has increased in recent years, reflecting growing awareness of perinatal substance use and its impact on maternal and neonatal outcomes. This trend parallels rising substance use rates across the general and perinatal populations, underscoring the urgency of addressing provider-level stigma as a barrier to compassionate, effective care.

Intervention strategies were notably non-standardized, with studies employing a wide range of formats, including didactic modules, workshops, clinical immersion, and arts-based approaches. While this diversity allows for contextual adaptation, it also limits comparability across studies and challenges efforts to identify core elements of effective training. Similarly, evaluation methods varied widely, with inconsistent use of validated measures, limited follow-up time points, and few studies assessing sustained behavior change. Despite this heterogeneity, most studies reported positive outcomes, including increased knowledge, improved provider confidence, and reduced stigmatizing attitudes. Interventions that incorporated real patient narratives, individuals with lived experience, or immersive settings often showed stronger impacts on empathy and attitude shift. However, fewer studies demonstrated long term changes in clinical practice, such as routine screening or reductions in punitive reporting behaviors. This limitation may be due to the nature of pre-post testing and the difficulty with sustained follow up, as they did not show *lack of long term efficacy*, but instead *were not able to measure long term efficacy*. This suggests a need for long term follow up and continued reinforcement and systems-level alignment.

Looking ahead, it is essential to prioritize accessible and equitable training, particularly for providers in high-burden or under-resourced settings. Many patients with perinatal SUDs receive care outside of large health systems or better-resourced metropolitan areas, where SUD training and access to other treatment resources may be scarce. Given the clear links between perinatal SUDs, mental health, and adverse obstetric outcomes, provider education is a clinical and public health necessity, not just a professional development opportunity. Future research should aim to develop standardized outcome measures, test long-term effectiveness, and integrate lived experience in training design. Scalable, sustainable formats, such as hybrid learning and quality improvement initiatives, may enhance reach and implementation. As the healthcare education field evolves, collaborative, interdisciplinary efforts will be key to advancing stigma-free, person-centered perinatal care.

## Data Availability

Data can be made available upon reasonable request to the authors.

